## Supplementary file 1 for "Exploring current and potential roles of informal healthcare providers in tuberculosis care in West Bengal, India: a qualitative content analysis"

**Table 1: Roles of IPs in TB care (classified based on TB care functions)**

| **Types of care** |  | **IPs TB care roles (by functions) *** | **Definitions** |  | **Refinements of roles based on the qualitative study findings **** | **Definitions** |
| --- | --- | --- | --- | --- | --- | --- |
| **Prevention** | 1 | Health promotion and education | Services include (but are not limited to) awareness-raising and social mobilization activities. | 1 | Health promotion and education | Activities related to promoting information on TB in the community or providing health education to any patient visiting IPs clinic before being diagnosed as having TB. |
|  | 2 | Immunization | Activities related to BCG Vaccination | 2 | Immunization | Activities related to BCG Vaccination |
|  | 3 | Latent TB infection screening | Activities related to screening of latent tuberculosis | 3 | Latent TB infection screening | Activities related to screening of latent tuberculosis |
|  | 4 | Latent TB infection prescription | Activities related to prescription of preventative treatment | 4 | Latent TB infection prescription | Activities related to prescription of preventative treatment |
|  | 5 | Latent TB infection administration | Activities related to administration of preventative treatment | 5 | Latent TB infection administration | Activities related to administration of preventative treatment |
| **Detection and diagnosis** | 6 | Active case finding | Activities related to active finding of TB cases in any setting | 6 | Active case finding | Activities related to active finding of TB cases in any setting |
|  | 7 | Passive case finding and referral | Activities related to screening and referral of TB cases among patients who visit IPs clinic | 7 | Passive case finding and referral | Activities related to screening and referral of TB cases among patients who visit IPs clinic |
|  |  |  |  | 8 | Accompany suspected TB cases to a health facility | Activities related to accompanying a suspected case of TB to a health facility |
|  | 8 | Clinical evaluation-TB | Activities related to clinical evaluation of TB among suspected cases | 9 | Clinical evaluation-TB | Activities related to clinical evaluation of TB among suspected cases |
|  | 9 | Laboratory examination and/or X-ray | Activities related to laboratory examination of TB specimens and X-ray examination | 10 | Collection and transportation of sputum samples | Activities related to collection and transportation of sputum samples to designated facilities |
|  |  |  |  | 11 | Contact tracing | Activities related to screening of contacts who were exposed to a case of confirmed TB |
| **Treatment and support** | 10 | Treatment initiation | Activities related to prescription of TB drugs to confirmed patients | 12 | Treatment initiation | Activities related to prescription of TB drugs to confirmed patients |
|  | 11 | Treatment supporter | Activities related to supporting TB treatment | 13 | Treatment supporter | Activities related to supporting TB treatment |
|  | 12 | Monitoring treatment progress and response | Activities related to periodic clinical evaluation and lab monitoring | 14 | Monitoring treatment progress and response | Activities related to periodic clinical evaluation and lab monitoring |
|  | 13 | Prevention and detection of adverse events and comorbidities | Activities related to monitoring of adverse drug reactions | 15 | Prevention and detection of adverse events and comorbidities | Activities related to monitoring of adverse drug reactions |
|  | 14 | Diagnosis and treatment of adverse events and comorbidities | Activities related to treatment of adverse drug reactions | 16 | Diagnosis and treatment of adverse events and comorbidities | Activities related to treatment of adverse drug reactions |
|  | 15 | Treatment lab monitoring | Activities related to conduct of laboratory examinations during treatment | 17 | Treatment lab monitoring | Activities related to conduct of laboratory examinations during treatment |
|  | 16 | Counselling and psychological support | Activities related to providing counselling and psychological support during treatment | 18 | Counselling – During TB treatment | Activities related to providing counselling and psychological support during treatment |
|  | 17 | Social support | Activities related to social support for a TB patient | 19 | Social support | Activities related to social support for a TB patient |

*IPs’ roles identified in the scoping review

**IPs’ roles refined based on the qualitative study findings
