## Supplementary file 2 for "Exploring current and potential roles of informal healthcare providers in tuberculosis care in West Bengal, India: a qualitative content analysis"

**Interview Guide for Informal Healthcare Providers:**

Icebreaker Question:

1. Tell me about your work as an IP in the community.

Probes:

- What kind of services do you provide?

2. Can you please share your current work with TB patients? Let them express first.
3. Can you please share your interaction with the last TB patient?

Probes:

- Ask how their experience was providing care to that patient. Ask questions like why they felt it was a good or bad experience.
- What kind of TB care services were offered?
- Ask if they encountered any challenges while providing TB care to that patient or any patient. Ask for details about the challenges. Examples include feeling like not having enough knowledge or did not know what to do with that case.

4. In your opinion, what kind of work/task can IPs do in TB care?

First, let IPs respond to this question. But if they are not able to provide a response, probe by asking about the following roles in TB care.

Probes:

- Creating community awareness on TB
- Door-to-door identification of TB patients
- Referring suspected TB patients from their clinic to government health centers
- Collecting and transporting sputum samples to the laboratory
- Initiating treatment for new TB patients
- Treatment supporter
- Providing counselling to TB patients when on treatment
- Follow up of patients lost to treatment
- Any other roles they think can be undertaken by IPs in TB care

Ask how and why questions, like how can they refer patients, would there be any challenges to undertaking these roles? What kind of support will they need from the government? If they think they cannot do those specific roles, ask for more details.

5. Do you think IPs have the knowledge and skills to undertake the roles that you mentioned above?

Probes:

- If yes, why do you think so? If not, what can help IPs to improve their knowledge and skills?

6. Is there any other information that you would like to share with me?

Thank you for your participation!!!

**Interview guide for formal providers:**

Icebreaker Questions:

1. Can you please share your work in TB care?

Probes:

- What is your major/primary role in the TB care program?

Now, I will focus our discussion on Informal Providers, who are locally known as RHCPs (Rural healthcare providers) in West Bengal.

2. Can you describe any experience you have working with an IP in general or in any TB program? Or have you had any chance to interact with IPs in general or any TB-related activities?
3. In particular, how are IPs providing care to TB patients in the community, in your opinion?

Probes:

- What kind of services are they providing to the patients?
- Do you think they have the appropriate knowledge to undertake those activities?

4. A significant proportion of TB patients seek care from IPs, or IPs are the first point of contact. Why do you think such practices exist in the community?
5. If IPs are engaged in the TB program, what do you think is the maximum level of the health system in which they can be involved?
6. In your opinion, what kind of activities, roles, or functions can they do in TB care?

Probes:

- Screening of TB symptomatic
- Referral of TB patients
- Diagnosis of TB patients
- Providing DOTs
- Counselling of patients
- Follow-up of patients
- We also did interviews with IPs in this study, and some of them expressed that they want to provide services such as referral of symptomatic cases, collect sputum samples, and be a treatment supporter. What is your opinion on that?

7. Is there any other information that you would like to share with me?

Thank you for your participation!!!
